# Ten-years absolute risk estimates of death and kidney failure in adults with chronic kidney disease: Analysis of electronic health records of 142,770 patients of the Social Security system of Peru

**DOI:** 10.64898/2026.06.18.26355937

**Authors:** Jessica Ivonne Bravo-Zúñiga, Winnie Contreras-Marmolejo, Obert Marín-Sánchez, Percy Soto-Becerra, Edgar Juan Coila-Paricahua, Isabel Álamo-Palomino, Madelaine Huanca-Roca, Lizbeth Arce-Gallo, Luis Randy Loayza-Arroyo, Mitshell Bikram Ramos-Quispe, Bryan Christopher Bastidas-Reyes, Daysi Zulema Díaz-Obregón

## Abstract

**Objective:** To determine the absolute risk of starting dialysis versus mortality among adults with chronic kidney disease (CKD) treated at EsSalud from 2013 to 2022, utilizing data from the Renal Health Surveillance system (VISARE).

**Methods:** This retrospective cohort study analyzed clinical records from the VISARE system (EsSalud). We estimated rates of dialysis initiation and death using Fine & Gray competitive risk models. Additionally, we calculated Restricted Mean Survival Time (RMST), adjusting for age, sex, clinical stage, and geographic region.

**Results:** Among 142,770 adults with confirmed CKD and available glomerular filtration rate data, only 15.2% had albumin-to-creatinine ratio measurements, allowing KDIGO staging of 40,404 patients (28.3%). Mortality without having previously started dialysis exceeded the probability of starting renal replacement therapy (RRT) from G1, becoming more marked in G3 of chronic kidney disease (CKD); the possibility of dialysis is only greater, as expected, in G5.

This outcome was most prevalent in regions with limited healthcare coverage. The combination of diabetes, hypertension, and age over 55 (the “triad”) was associated with reduced restricted mean survival time at both 5- and 10-year horizons across all enrollment cohorts. While Lima saw the highest rates of renal replacement therapy initiation, the Andean and Amazonian regions reported the lowest indicators.

**Conclusions:** Death without prior dialysis was the dominant outcome from G1 to G3 in this Peruvian cohort with national insurance, with direct implications for prognostic counseling, recalibration of renal failure risk equations, and equitable expansion of nephrology services in underserved regions.

## Introduction

Chronic kidney disease (CKD) is defined by a progressive and irreversible decline in kidney function. Frequently asymptomatic in its early stages, CKD often remains undetected until advanced disease, when cardiovascular and metabolic complications become clinically evident (1,2). The global prevalence of CKD is estimated at 11%–13%, with stage 3 as the most common presentation(3). Although CKD shares traditional risk factors with cardiovascular disease (CVD)—including hypertension, diabetes mellitus, and dyslipidemia (4)—it also involves distinct pathophysiological pathways. Endothelial dysfunction, vascular calcification, and activation of the renin–angiotensin–aldosterone system contribute to excess cardiovascular risk, even in early CKD(5–8).

In Latin America, CKD has increased substantially in recent decades and is now a major contributor to mortality and disability-adjusted life years (9,10), driven largely by population aging and the rising burden of non-communicable diseases (11). In Peru, CKD represents a significant public health challenge, compounded by delayed diagnosis and limited access to renal replacement therapy (RRT), including hemodialysis(12). These constraints amplify disease burden, particularly in settings with fragmented healthcare delivery. The COVID-19 pandemic exacerbated these structural challenges: some studies document an increase in hospitalizations and mortality in Peruvian patients with chronic dialysis during 2020-2021(13).

In CKD, a “competing risk” occurs when an event—such as death—prevents the occurrence of another event, like starting dialysis. Because older patients or those with severe comorbidities are highly likely to die before their kidneys fail, ignoring this leads to a severe overestimation of dialysis(14).

Long-term follow-up is essential, as CKD progression typically takes years or decades; therefore, longitudinal studies allow for the calculation of true incidence rates of these serious outcomes (death and need for dialysis). In addition, there is a lack of local data: most of the evidence comes from high-income countries (Chronic Renal Insufficiency Cohort (CRIC), CKD Prognostic Consortium)(15). In Latin America, there is heterogeneity (genetic, socioeconomic, and environmental) that affects disease progression and outcomes. Therefore, regional registries show an increase in the prevalence/incidence of end-stage renal disease but lack detailed follow-up from the initial stages.(16).

The results will have an impact on public health policies, allowing for the estimation of future burden (need for dialysis stations, system costs), prioritization of primary and secondary prevention (control of risk factors, use of SGLT2 inhibitors, ACE inhibitors/ARBs), improved equity, and the promotion of resource allocation. In conclusion, this study will contribute to addressing knowledge gaps (unmet needs)(17).

This study aims to estimate the 10-years absolute risks of dialysis initiation and mortality among adults with CKD treated within the EsSalud system between 2013 and 2022, using data from the Renal Health Surveillance System (VISARE). We also estimate restricted mean survival time (RMST) as a prognostic measure within a competing risks framework adjusted for age, sex, CKD stage, and geographic region.

## Methods

### Study Design

A retrospective cohort study was conducted to describe the prognosis of the following relevant clinical outcomes: dialysis initiation and mortality in adult patients with chronic kidney disease (CKD). The study analyzed clinical records from the EsSalud Renal Health Surveillance System (VISARE), corresponding to the period between January 2013 and December 31, 2022. These data were originally collected in the context of the external validation study of the Kidney Failure Risk Equation (KFRE)(18), which used a subset of patients with stage G3-G4 CKD.

For the present analysis, which includes the entire patient cohort, new authorization was obtained from the Research Ethics Committee of the Edgardo Rebagliati National Hospital of EsSalud (Code No. 23-GRPR-ESSALUD-2025, approved on January 24, 2025). The research follows a type 1 forecasting framework in accordance with the PROGRESS guidelines (19) and adheres to the STROBE statement (Strengthening the reporting of observational studies in epidemiology) for observational research (20,21).

Data extraction, linkage, and processing were performed solely by one investigator (EJC-P), who was a study researcher at IETSI, EsSalud, with authorized access to the raw data sources as part of the institution’s renal health surveillance operations. The remaining authors did not have access to information that could identify individual participants during or after data collection.

### Data Sources and Study Population

The data were integrated from three primary sources: (1) the VISARE subsystem, through manual clinical records from 2013 to 2022; accessed on August 25, 2023; (2) from the National Center for Renal Health and Hospital Management System (SGH) through the identification of the dialysis procedure record, accessed on September 8, 2023; (3) data from the Central Insurance Management Office, which constantly receives vital information from RENIEC (National Registry of Identification and Civil Status), accessed on August 2, 2023.

The study population included adults (≥18 years) with a confirmed CKD diagnosis based on the Estimated Glomerular Filtration Rate (eGFR) ml/min per 1.73 m² according to the 2021 CKD-EPI equation(19) and the albumin-to-creatinine ratio (ACR) in mg/g was used for stages 1 and 2. Cases lacking diagnostic confirmation were excluded.

### Outcome and Variables of stratification

The primary outcomes analyzed were:

- **Dialysis initiation:** This variable required a combination of multiple data sources and criteria. The main proxy for kidney failure was the exact date of the first hemodialysis, obtained from the digital biological record of the National Center for Renal Health, which maintains the national dialysis data for EsSalud. We also cross-referenced this information with the electronic health records of the Hospital Management System (HMS) since 2013. Renal insufficiency was identified if patients had any of the following ICD-10 codes in their records:

`o` N18.5 (chronic kidney disease, stage 5)
`o` N18.6 (End-stage renal disease)
`o` Z99.2 (Dependence on renal dialysis)
`o` Z49.1 (Extracorporeal dialysis)
`o` Z49.2 (Other specified dialysis)
`o` Z94.0 (Kidney transplantation) This computational phenotyping method, based on ICD-10 coding, follows a previously established scheme used in the literature (3).
- **Mortality:** Mortality: overall time to death and death before dialysis. Mortality data were obtained by linking information from two national sources: the Peruvian National Death Registry (SINADEF) and the National Registry of Identification and Civil Status (RENIEC), both accessible through the Office of Insured Services. This approach provided a comprehensive and accurate method for identifying the time to kidney failure in patients, considering the healthcare context and practices within EsSalud.

Key explanatory variables included age, sex, macro-region of origin, hypertension, diabetes mellitus, and clinical CKD stage.

### Statistical Analysis

Descriptive analyses were stratified by clinical stage and region. Five- and ten-year cumulative incidence rates for death and dialysis initiation were estimated using Cumulative Incidence Functions (CIF), accounting for the competing nature of these outcomes. For the analysis of dialysis initiation, death without prior dialysis was considered as a competing event. Furthermore, Restricted Mean Survival Time (RMST) at 5 and 10 years was calculated using flexible parametric survival models. In the presence of competing risk, CIF were estimated using the non-parametric Aalen–Johansen estimator, which provided unbiased estimation of event probabilities when the competing events preclude the occurrence of the outcome (20,21). RMST estimates were stratified by demographic and clinical characteristics. Ninety-five-percent confidence intervals for RMST were obtained by non-parametric percentile bootstrap with B = 1000 resamples (seed = 1234). Statistical analyses were performed in R version 4.3 (R Core Team, 2024).

## Results

Between 2013 and 2022, 148,277 patients were registered in the National Renal Health Surveillance Registry (VISARE). Of these, 143,202 were identified as adults with risk factors (diabetes mellitus, hypertension, and/or age over 55). Of these, 142,770 (99.7%) had complete data for calculating the estimated glomerular filtration rate (eGFR), while only 21,792 (15.2%) had an albumin-to-creatinine ratio (ACR) available (Figure 1).

**Figure 1.**
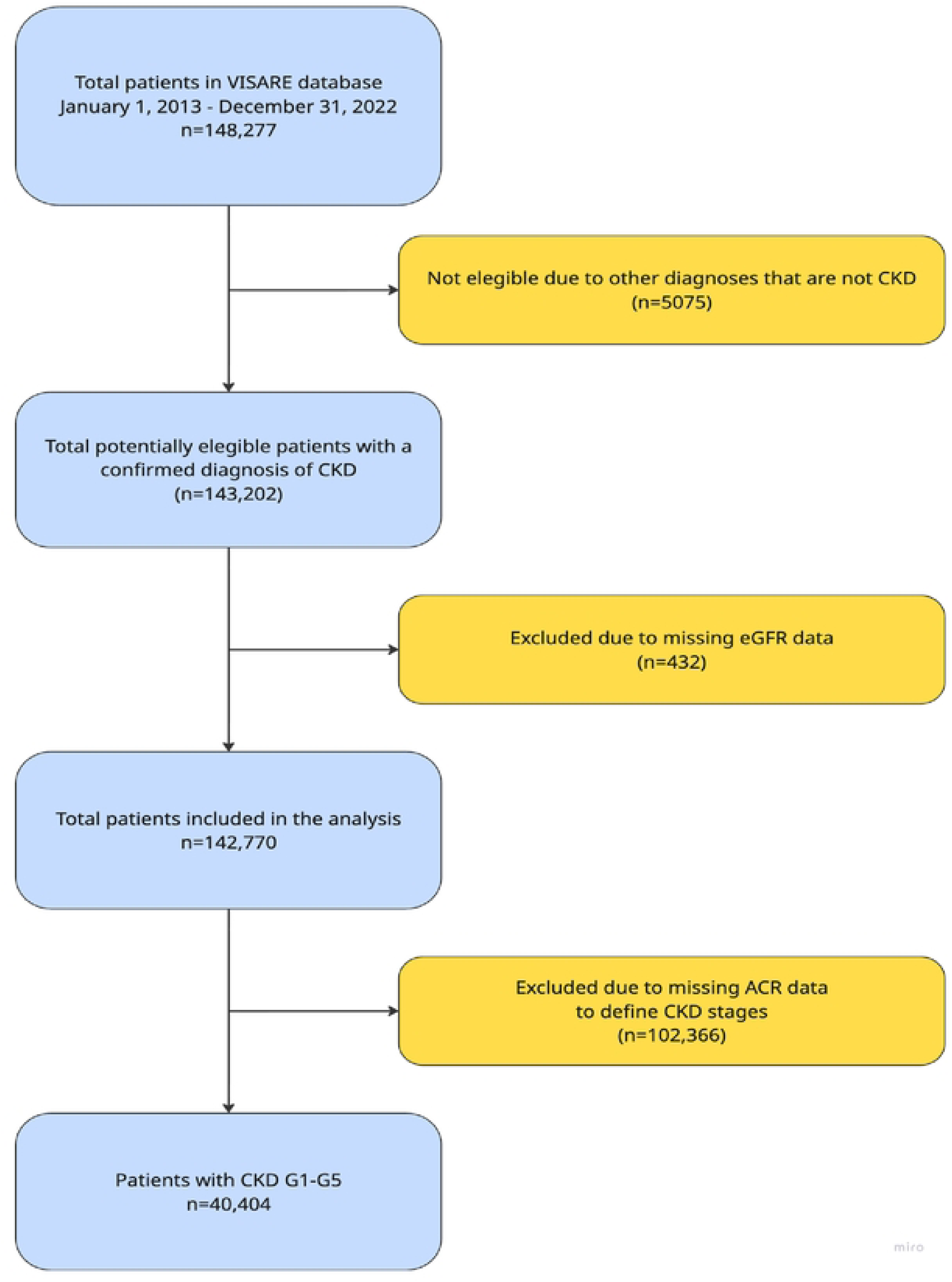
Flow diagram of included patients. GFR, glomerular filtration rate; CKD, chronic kidney disease; ACR, albumin-to-creatinine ratio

The cohort was predominantly female (53.8%), with over 70% being older adults (median age: 66 years [IQR 58-74]). Geographically, the largest proportion of patients (37.2%) were from Lima, while the Amazon region (eastern Peru) accounted for only 7.1% of patients. The baseline characteristics of these patients are presented in Table S1. When compliance with the KDIGO (kidney disease: Improving Global Outcomes) guidelines criteria was assessed, 40,404 (28.2%) patients were classified as having chronic kidney disease (CKD) (Figure 2). Table 1 summarizes the clinical staging of CKD patients stratified by sex, age, comorbidities (diabetes mellitus [DM] and hypertension [HTN]), and the cumulative incidence of dialysis and mortality. A female predominance was observed in all stages, except for G3b, where men predominated (53.0%). Age showed a direct correlation with CKD progression, except in stage G5, where the mean age was slightly lower than in the intermediate stages (67.1 years). HTN affected 24,211 (69.4%) of the patients evaluated, while 11,459 (35.9%) patients had DM. Regarding clinical outcomes, 2477 (6.1%) patients-initiated dialysis during the follow-up period. This progression to dialysis was directly related to the stage of CKD, being most prevalent in stages 4 (31.3%) and 5 (54.5%). Similarly, mortality increased progressively from 10.6% in stage G1 to 52.5% in stage G5. This trend highlights the increasing burden of complications associated with the progression of renal impairment.

**Figure 2:**
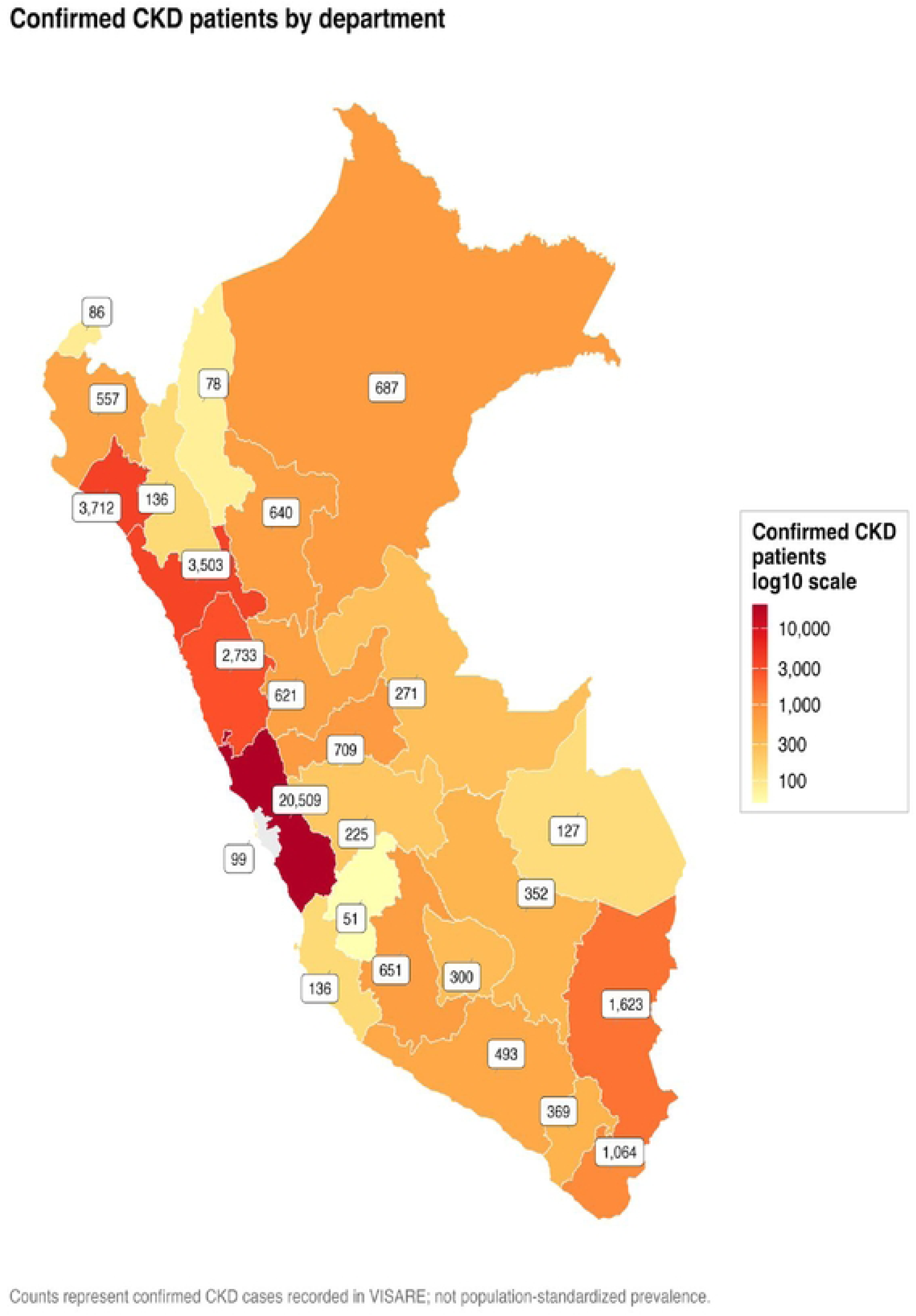
Map of patients with chronic kidney disease included by department of the National Renal Health Surveillance Registry (VISARE) of EsSalud, 2013-2022

**Table 1:**
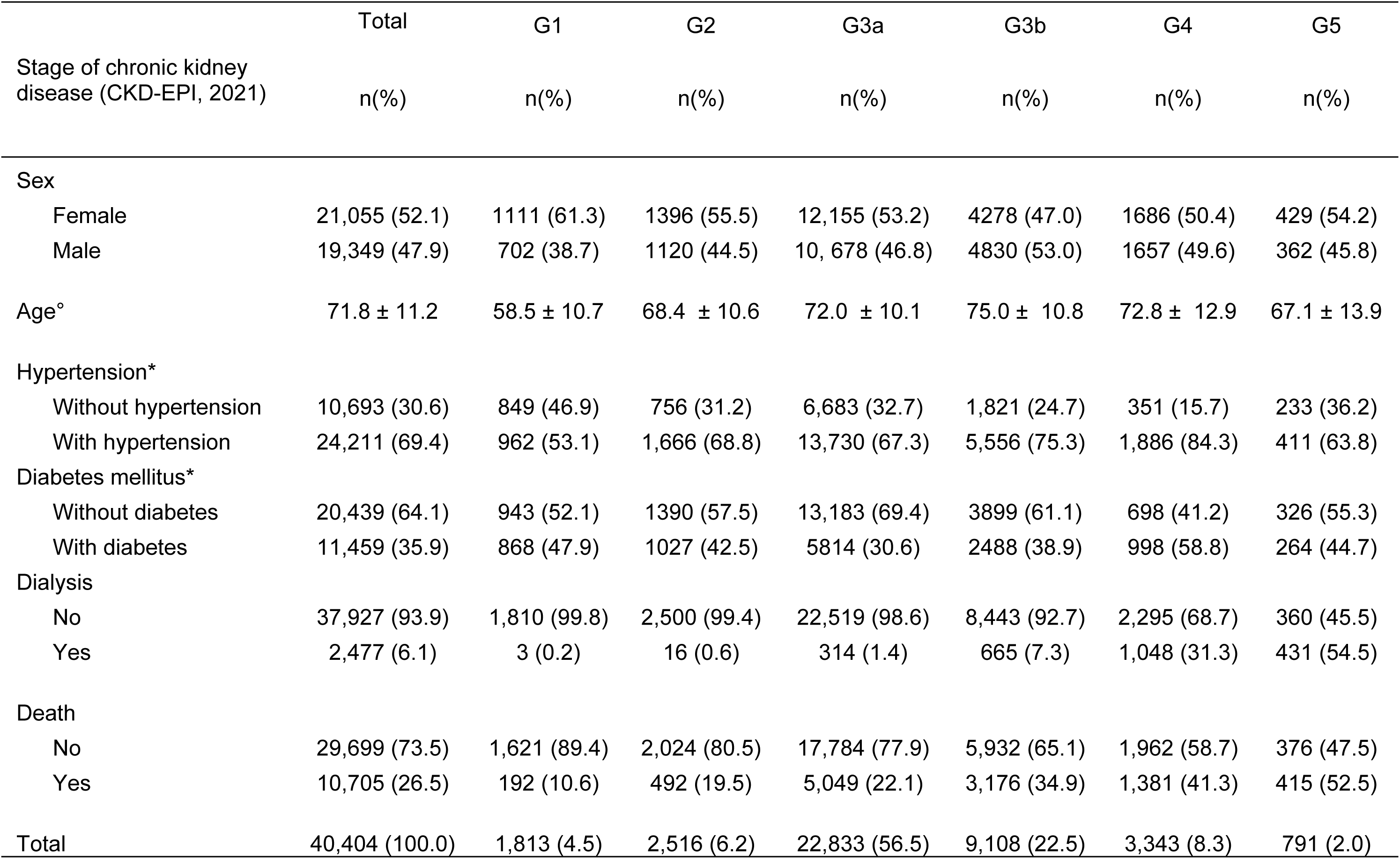
Sociodemographic and clinical characteristics of patients according to their clinical stage of chronic kidney disease, National Renal Health Surveillance Registry (VISARE) of EsSalud, 2013-2022.

### Clinical Outcomes: Dialysis and Mortality

In the general population, the requirement for renal replacement therapy (RRT) rose sharply in stage G4 (25.8%) compared to earlier stages (range: 0.2%–6.0%). For patients with co-occurring DM and HTN, the requirement for RRT in stage G4 was even higher than in those who did not have these conditions (28.6%). The risk of death increased progressively with disease stage from stage 1 to 3b (range: 10.4%–31.4%). Mortality was notably higher among patients with diabetes, hypertension, and those over 55 years of age (range: 19.2%–37%) (Table 2). The cumulative incidence of dialysis increases progressively as kidney disease progresses, however patients in advanced stages (G4 and G5) showed a pronounced increase in dialysis initiation, reaching 25.8(24.3-27.4) and 44.4(40.6-48.3) respectively, while G1 and G2 remained nearly flat. This clear separation between curves aligns with the pathophysiology of CKD, where accelerated functional loss in late stages necessitates earlier RRT intervention. (Figure S1). The geographical analysis of dialysis initiation revealed marked territorial inequality. Lima and Callao exhibited the highest cumulative probability of starting RRT. In contrast, other macro-regions showed nearly flat incidence curves, suggesting limited access to or under-utilization of dialysis outside the capital.

**Table 2.** Dialysis and five-year mortality incidence according to stage of chronic kidney disease, National Renal Health Surveillance Registry(VISARE)

| Stage of chronic kidney disease (CKD-EPI) | Total (%)<br>[IC 95%] | Diabetes mellitus (DM) (%)<br>[IC 95%] | Hypertension (HT) (%)<br>[IC 95%] | DM+HTA (%)<br>[IC 95%] | Age >55 (%)<br>[IC 95%] | DM+HTA+Age>55 (%)<br>[IC 95%] |
| --- | --- | --- | --- | --- | --- | --- |
| **Dialysis** |  |  |  |  |  |  |
| G1 | 0.2 (0.1-0.5) | 0.3 (0.1-1.0) | 0.0 (0.0-0.4) | 0.0 (0.0-0.8) | 0.2 (0.0-0.6) | 0.0 (0.0-1.1) |
| G2 | 0.5 (0.3-0.8) | 0.7 (0.3-1.4) | 0.5 (0.3-1.0) | 0.9 (0.4-1.9) | 0.5 (0.2-0.8) | 0.7 (0.3-1.7) |
| G3a | 1.0 (0.9-1.2) | 1.7 (1.4-2.1) | 1.0 (0.9-1.2) | 1.9 (1.5-2.4) | 0.9 (0.8-1.1) | 1.8 (1.4-2.3) |
| G3b | 6.0 (5.5-6.5) | 8.7 (7.7-9.9) | 5.8 (5.2-6.4) | 8.8 (7.6-10.2) | 5.4 (5.0-5.9) | 7.9 (6.7-9.2) |
| G4 | 25.8 (24.3-27.4) | 29.5 (26.6-32.5) | 24.4 (22.5-26.5) | 28.6 (25.4-32.0) | 25.1 (23.5-26.7) | 27.8 (24.6-31.2) |
| G5 | 44.4 (40.6-48.3) | 41.7 (35.4-48.4) | 43.4 (38.2-48.6) | 43.4 (36.1-51.0) | 44.2 (40.0-48.4) | 42.3 (34.4-50.5) |
| **Death** |  |  |  |  |  |  |
| G1 | 10.4 (9.1-11.9) | 13.6 (11.5-16.0) | 12.0 (10.1-14.2) | 16.9 (13.8-20.5) | 13.2 (11.4-15.3) | 19.2 (15.3-23.7) |
| G2 | 19.0 (17.5-20.6) | 22.9 (20.4-25.5) | 21.5 (19.6-23.6) | 24.9 (21.9-28.1) | 20.9 (19.3-22.6) | 26.7 (23.5-30.1) |
| G3a | 21.4 (20.9-21.9) | 25.5 (24.4-26.6) | 23.2 (22.5-24.0) | 27.4 (26.1-28.8) | 22.2 (21.7-22.8) | 28.1 (26.7-29.5) |
| G3b | 31.4 (30.5-32.4) | 34.3 (32.5-36.2) | 33.4 (32.1-34.6) | 36.2 (34.1-38.4) | 32.3 (31.3-33.3) | 37.0 (34.8-39.2) |
| G4 | 30.2 (28.6-31.9) | 31.5 (28.6-34.6) | 31.7 (29.6-33.9) | 32.8 (29.5-36.3) | 31.4 (29.8-33.2) | 34.0 (30.6-37.6) |
| G5 | 27.2 (23.9-30.8) | 31.7 (25.8-38.1) | 27.5 (23.0-32.4) | 30.1 (23.7-37.5) | 29.1 (25.4-33.1) | 31.7 (24.6-39.7) |

### Competing Risks: Dialysis vs. Mortality

This analysis included 39,732 patients who met the follow-up time requirement. The cumulative incidence of death without dialysis remained consistently higher than the cumulative incidence of dialysis initiation over a five-year follow-up (Figure 3). By year five, the cumulative incidence of death reached approximately 18%, whereas dialysis initiation reached about 5%. These findings suggest that death is a more frequent outcome than progression to RRT in the overall CKD population.

**Figure 3.**
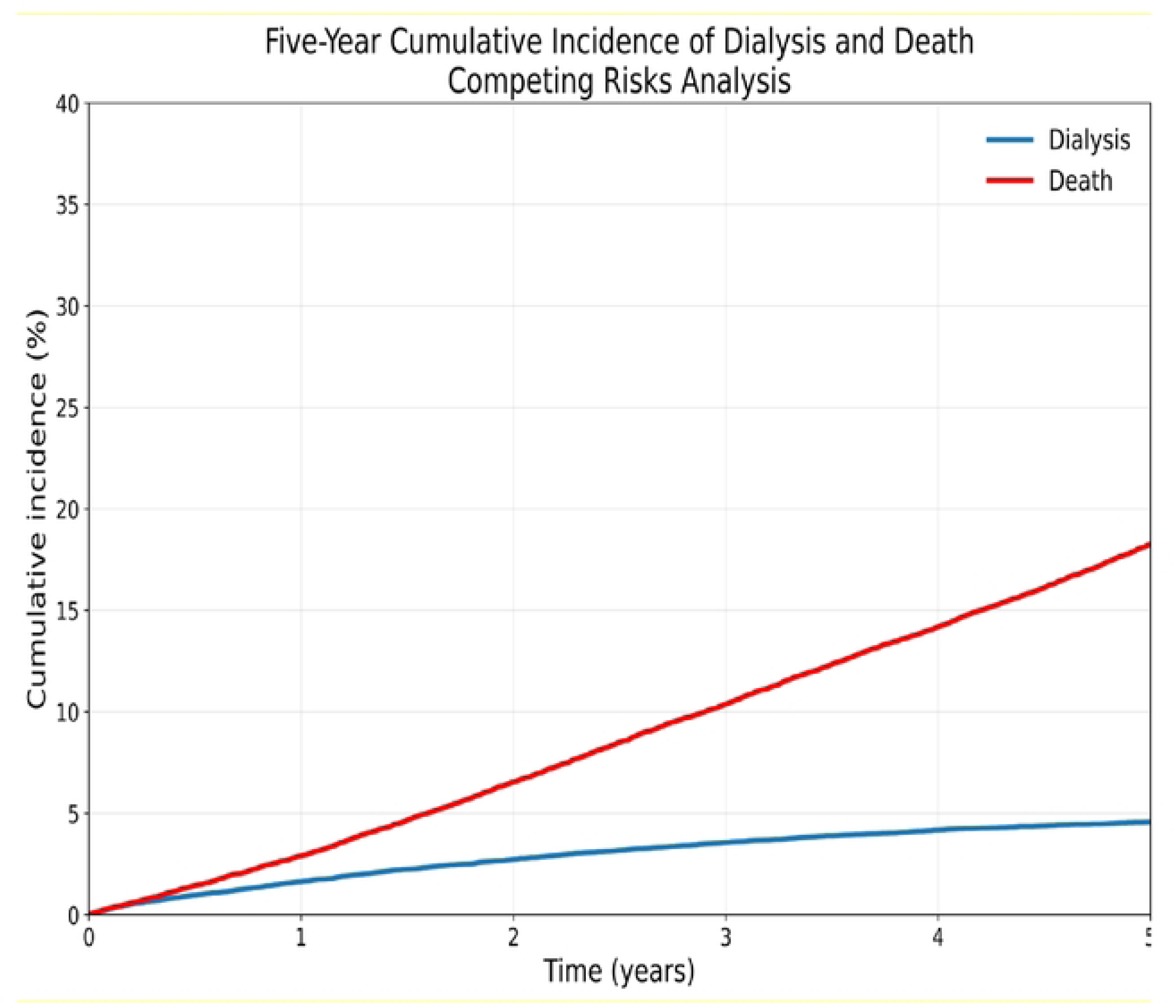
Analysis of competing risks between dialysis initiation and death without dialysis over five years of patients from the National Renal Health Surveillance Registry (VISARE) of EsSalud, 2013-2022

Stratification by CKD stage revealed marked differences in the relative occurrence of both outcomes. In stage G1 through G3b, death remained substantially more frequent than dialysis initiation. In stage G4, the cumulative incidences of death and dialysis became similar over time. However, in stage G5, dialysis initiation exceeded mortality, reflecting the increasing likelihood of RRT among patients with advanced kidney disease (Figure 4).

**Figure 4.**
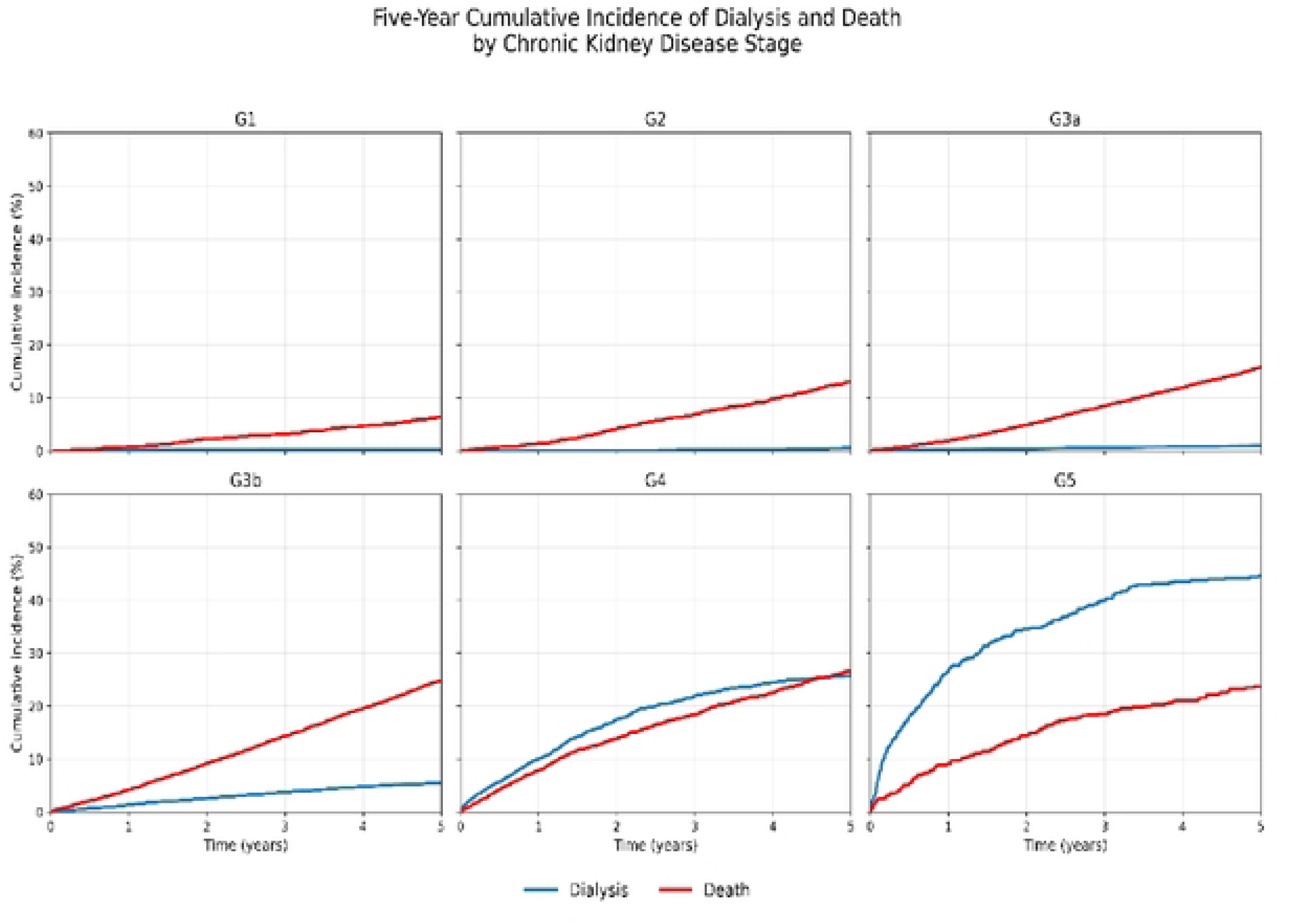
Analysis of competing risks between dialysis initiation and death without dialysis by stages of chronic kidney disease over five years of patients from the National Renal Health Surveillance Registry (VISARE) of EsSalud, 2013-2022

In contrast, among younger adults (≤55 years) without diabetes or hypertension (Figure S2), the competing risk pattern differed from the observed in the overall population. In stages G3b and G4, the cumulative incidence of dialysis initiation was like or slightly higher than the incidence of death. In stage G5, dialysis initiation clearly exceeded mortality throughout follow-up.

### Restricted Mean Survival Time (RMST)

The figure 5, show RMST (Restricted Mean Survival Time) at 5 and 10 years according to the year of enrollment and stratified by the number of conditions present in the triad: DM, HTN and Age >55 years.

**Figure 5:**
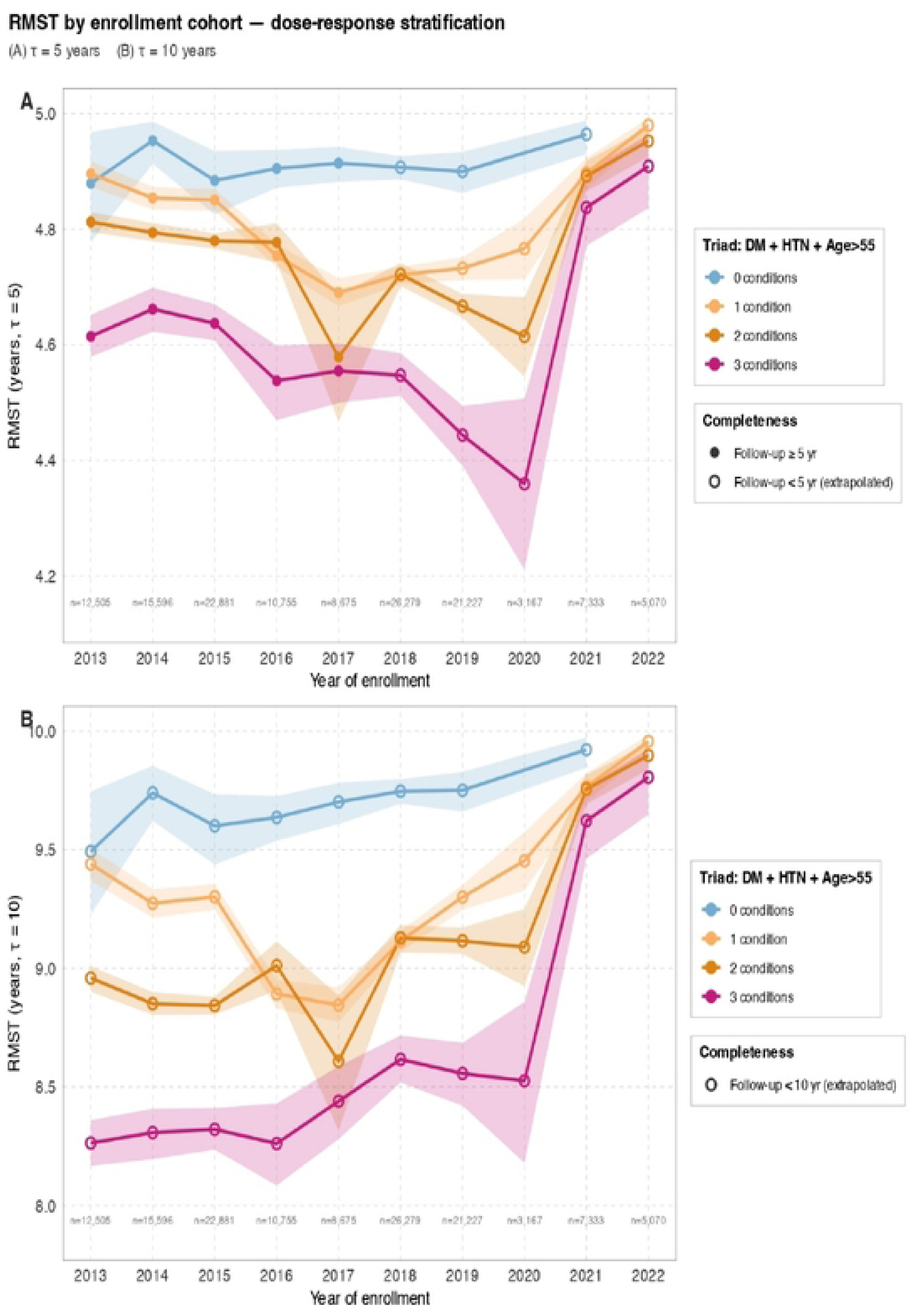
Restricted Mean Survival Time (RMST) at 5 and 10 years according to comorbidity

The RMST is truncated at τ = 5 and 10 years, so it can be interpreted as the average number of years lived/free of the event during the first 5 o 10 years of follow-up. There is a clear prognostic gradient in patients without major comorbidities; 1 condition: intermediate RMST, 2 conditions: lower RMST and 3 conditions, worse RMST.

This suggests a strong cumulative risk burden effect. Subjects without comorbidities maintain an approximate RMST of 9.5–9.9 years, while subjects with all three conditions maintain an RMST of 8.2–9.8 years, depending on the cohort. The maximum difference between extremes is approximately ΔRMST of 1.3–1.5 years. Furthermore, the line of patients without comorbidities maintains low interannual variability, relatively narrow confidence intervals, and consistently high RMST. On the other hand, the group with three conditions has the worst prognosis and greatest instability, with wider confidence bands and abrupt changes between 2020 and 2021, coinciding with the SARS-CoV-2 pandemic. This suggests a synergy between diabetes, hypertension, and aging.

## Discussion

In this national cohort of patients insured by EsSalud, mortality without having previously started dialysis exceeded the probability of starting renal replacement therapy (RRT) from G1, becoming more marked in G3 of CKD; the possibility of dialysis is only greater, as expected, in G5.

These findings highlight a dominant competing risk of death that fundamentally shapes the evolution of CKD in this context(22). The clinical implication is clear: failing to account for competing mortality can bias risk estimation and decision-making. Widely used tools, such as the Kidney Failure Risk Equation (KFRE), do not explicitly model death as a competing event; censoring for mortality leaves the estimated risk of kidney failure unchanged and, therefore, may overestimate progression to end-stage renal disease (ESRD). This limitation is particularly relevant in populations with a high cardiovascular burden, where death often precedes ESRD (22,23). In line with this, previous validation of the KFRE formula in EsSalud demonstrated incorrect calibration, necessitating a second study in which recalibration was performed to include competing risk. Interestingly, when the risk-utility curve is applied, the recalibrated and original formulas provide similar clinical benefit and could therefore be used interchangeably(18,24).

These patterns are plausible from a biological and epidemiological perspective. CKD in Peru is primarily due to diabetes mellitus and hypertension (25,26), conditions strongly associated with cardiovascular mortality. As kidney function declines, cardiovascular risk accelerates, tipping the balance toward death rather than progression to end-stage renal disease (ESRD). In contrast, younger individuals without major metabolic comorbidities showed slower progression and a higher likelihood of surviving to renal replacement therapy (RRT), suggesting that patients who start dialysis at advanced stages may represent a select subgroup with more favorable risk profiles: the survivors.

Healthcare system factors are likely to amplify this competing risk structure. Limited access to renal replacement therapy (RRT), delayed referrals, and fragmentation between primary and specialized care reduce the likelihood of timely dialysis initiation, particularly outside of Lima. (27,28)

The finding that only 40% of patients with stage G5 CKD initiate dialysis raises the question of what happened to the remaining patients. Was this due to individual characteristics or is it consistent with the relatively low coverage of renal replacement therapy (RRT) in Peru compared to Latin America (12,25). Taken together, these results suggest that excess mortality reflects not only the biology of the disease but also structural barriers to care.(29) (30).

The COVID-19 pandemic further exposed the vulnerability of the system. Patients undergoing hemodialysis experienced disproportionate increases in hospitalizations and mortality(13) Therefore, apparent improvements in survival after 2019 should be interpreted with caution, as they likely reflect selection effects, reduced access to renal replacement therapy, or delays in outcome determination, rather than genuine improvements in prognosis.

Geographic heterogeneity adds another layer of complexity. Evidence from the ISN-GKHA (International Society of Nephrology Global Kidney Health Atlas) indicates that the burden of chronic kidney disease in Latin America is determined by environmental and social factors(31). In Peru, structural inequalities in the Andean and Amazonian regions likely contribute to underdiagnosis, delays in care, and worse outcomes, reinforcing the need for geographically targeted strategies.

Another striking fact to consider is the low percentage of patients who had glomerular filtration rate (GFR) and albumin-to-creatinine ratio measurements. The gap between recommendations and actual practices is one of the most critical problems in preventive nephrology. While the KDIGO, ADA (American Diabetes Association) and AHA (American Heart Association) guidelines recommend annual ACR screening in at-risk populations, actual usage rates range from 4% to 35%(32) (33) (34) depending on the population studied and the country. A systematic review identified the greatest barrier to non-adherence to ACR screening guidelines as the perception that the test does not impact patient management, logistical and economic barriers, and the lack of quality incentives (35). With the expanding therapeutic arsenal for preventing complications of elevated albuminuria (including SGLT2 inhibitors), early identification and monitoring are more important than ever (36). However, ACR testing in the real world is low, particularly in non-diabetic patients with hypertension.

This study has limitations. The main one is the low percentage of patients with albuminuria data, which limits the diagnosis of CKD in early stages (stages I and II). It is possible that many patients evaluated solely by eGFR, with values above 60 ml/min, were classified as “healthy” due to the lack of evidence of kidney damage: albuminuria.

However, given that our primary outcomes were the initiation of dialysis and mortality, this limitation is unlikely to substantially alter the observed patterns of competing risk. Residual confounding factors and potential biases in registry-based data should also be considered.

Despite these limitations, the national scope of this analysis provides strong evidence that competing mortality is a dominant determinant of CKD outcomes in the social security-insured population of Peru.

These findings support a shift toward risk models that explicitly incorporate competing events and underscore the need to strengthen early detection, optimize multidisciplinary management of cardiovascular risk, and expand equitable access to renal replacement therapy.

## Conclusions

In this nationwide EsSalud cohort of 142,770 adults with chronic kidney disease followed between 2013 and 2022, Mortality without having previously started dialysis exceeded the probability of starting renal replacement therapy (RRT) from G1, becoming more marked in G3 of CKD, the possibility is equal in G4 and becomes greater as expected, in G5.

This evidence reveals a critical dynamic of competing risks, where progression to end-stage renal disease is prematurely interrupted by fatal outcomes associated with advanced age, a high burden of comorbidities, and structural barriers to accessing specialized care.

## Data Availability

In accordance with EsSalud's privacy policies, the minimum data set is not publicly accessible. However, we can provide the anonymized data upon reasonable request addressed to the corresponding author.

## Ethics statements

### Patient consent for publication

Not applicable.

### Ethics approval

This study was approved by the Research Ethics Committee of the Edgardo Rebagliati National Hospital at EsSalud (Code No. 23-GRPR-ESSALUD-2025) and was conducted in accordance with the principles of the Declaration of Helsinki. A waiver of informed consent was granted by the Research Ethics Committee of the Edgardo Rebagliati National Hospital at EsSalud because the study involved routinely collected secondary data. Given the minimal risks associated with this type of research, this waiver was deemed a reasonable justification and was approved accordingly.

## Notes

### Competing Interest Statement

The authors have declared no competing interest.

### Funding Statement

The author(s) received no specific funding for this work.

### Author Declarations

This study was approved by the Research Ethics Committee of the Edgardo Rebagliati National Hospital at EsSalud (Code No. 23-GRPR-ESSALUD-2025) and was conducted in accordance with the principles of the Declaration of Helsinki. A waiver of informed consent was granted by the Research Ethics Committee of the Edgardo Rebagliati National Hospital at EsSalud because the study involved routinely collected secondary data.

